# Analysis of health education demand for patients with HPV infection based on the KANO Model

**DOI:** 10.1101/2025.06.07.25329198

**Authors:** Yuying Zhang, Li Luo, Shuaihui Du, Yaling Zhang, Qian Guo, Lihua Zhou

## Abstract

**Objective:** To analyze the health education demand of patients with HPV infection based on the KANO model and provide a theoretical basis for the health education intervention of patients with HPV infection.

**Methods:** Participants were selected from West China Second University Hospital of Sichuan University in Chengdu, China. From June 1, 2024 to December 31, 2024, a cross-sectional survey was conducted to explore demand for the health education of patients with HPV infection, and data were analyzed based on the Kano model.

**Results:** A total of 428 patients were included. The results of this study show that the health education demand of patients with HPV infection include 23 one-dimensional demands and 5 attractive demands, The findings of the matrix analysis showed that 9 items were situated in the area of one-dimensional attributes quadrant, 5 items were situated in the area of attractive attributes quadrant, 11 items were situated in the area of indifferent attributes quadrant, and 3 items were situated in the area of must-be attributes quadrant.

**Conclusion:** Based on the Kano model, it was found that patients with HPV infection hold a positive attitude towards providing health education, and their demand for different health education services varies. These findings provide a theoretical basis for medical decision-makers to formulate health education, and also offer a scientific basis for nursing managers to improve health education services to meet the health education demand of patients with HPV infection.

## Introduction

Human papillomavirus (HPV) infection is closely related to the degree of human reproductive system lesions and the development of malignant tumors, especially high-risk HPV infection, which is an important cause of cervical cancer in women [1]. The infection rates of high-risk HPV16, 18, and 52 were relatively high among patients with different disease conditions (normal cervical cytology, precancerous lesions, and cervical cancer) [2–6].

In recent years, the development of medical technology and the deepening of treatment and prevention methods have increased the demand for health education services. People urgently need the popularization of knowledge related to HPV infection to effectively carry out prevention and treatment work. The "Comprehensive Prevention and Control Project for Cervical Cancer" also proposes that health education must run through the entire process [7]. Previous studies have shown that compared with 10 years ago, the awareness of cervical cancer, HPV and HPV vaccines among the Chinese population has increased. However, the relevant awareness is still limited. The good rate of knowledge about cervical cancer among women infected with HR-HPV is only 42.5% [8]. Some studies have shown that most women have expressed a positive attitude towards HPV screening. And it is required to know more about cervical cancer and its preventive measures [7]. Therefore, how to provide the health education that patients truly need is worthy of further exploration.

The KANO model is a simple and feasible technique for identifying service attributes. In the product management of enterprises, it classifies product attributes into five types based on the relationship between the objective performance of the product and the subjective feelings of customers: attractive quality, must-be quality, one-dimensional quality, indifferent quality and reverse quality, in order to formulate quality management strategies and improvement plans [9]. In recent years, the KANO model has also gradually been applied in the field of healthcare, such as health education for patients undergoing radiotherapy and chemotherapy for tumors [10], the research and development of intelligent medication systems for the elderly [11], the optimization of nursing service measures for children and their families [12], and the design of new health education platforms and models based on the Internet [13].

The KANO model provides a new idea for conducting health education for patients. Moreover, our research team has developed a questionnaire on the health education demand of HPV-infected patients under the KANO model and conducted a reliability and validity analysis[14]. Therefore, this study will analyze the health education service demand of HPV-infected patients and determine each demand attribute. Provide a basis for precise health education intervention for patients with HPV infection.

## Methods

### Sample and setting

From June 1, 2024 to December 31, 2024, patients with HPV infection who visited West China Second University Hospital of Sichuan University in Chengdu, China, were selected to make a survey. Inclusion criteria were as follows: (1) aged ≥16 years; (2) met the relevant diagnostic criteria for cervical lesions in Gynecology and Obstetrics, and were confirmed as cervical lesions by the pathological examination of Thinprepcy tologictest (TCT); (3)could communicate normally and cooperate with the investigation and evaluation; (4) Informed, voluntary participation in this study, signed informed consent. Exclusion criteria were as follows: (1) Patients with other malignant tumors; (2) with AIDS, syphilis and other infectious sexually transmitted diseases and with severe organ function injury; (3) with fungal vaginitis, bacterial vaginitis and other cervical inflammatory diseases.

According to the Kendall criterion, the sample size was 5 to 10 times the number of questionnaire items. The "KANO Model HPV Infection Patient Health Education demand Attribute Questionnaire" developed by our research team was used in this study, which had a total of 28 items. Considering a 20% loss to follow-up rate, the final sample size was determined to be 336 cases.

### Instruments

The investigation was conducted based on the "KANO Model Questionnaire on Health Education demand of HPV Infection Patients" formulated by Zhang’s[14] . Each item in this questionnaire consists of one positive question and one reverse question. The items are scored on a 5-level scale, representing "like (5 points)", "take for granted (4 points)", "don’t care (3 points)", "can tolerate (2 points)", "don’t like (1 point)". The higher the score, the greater the degree of approval.

Our initially developed Kano questionnaire had acceptable internal consistency, with standardized Cronbach’s alpha values of 0.940 and 0.955 for positive and reverse questionnaires, respectively. Kaiser–Meyer–Olkin (KMO) measures and Bartlett’s test of sphericity were used to test the validity, and the KMO values for the positive and reverse questionnaires were 0.950 and 0.965, respectively, which were greater than 0.7, and Bartlett’s test of Sphericity values of p<0.001, which indicated that the questionnaires had good validity[14].

### Data collection methods and quality control

In this study, the included patients were determined through the hospital’s big data system. With the help of the hospital’s follow-up platform, questionnaires were pushed through the hospital public account and short message. Two managers of the follow-up platform were first uniformly trained to modify the contents that needed to be adjusted in the online questionnaires and set the time for sending and receiving the questionnaires. The managers of the follow-up platform export data every month. For patients who have not completed the questionnaires, the system resend the questionnaires every week. For patients with missing items in the questionnaires, the researchers inquire about the results by phone and make supplements. Meanwhile, two staff members screened out questionnaires that took less than 5 minutes to fill out and had consistent answers and inconsistent answers. These questionnaires were regarded as invalid questionnaires. All patients signed written informed consent forms and completed questionnaires on the follow-up platform.

### Statistical Methods

The data were statistically analyzed by using SPSS22.0 software, and the demand attributes were analyzed by applying the Kano attribute classification method.

### Data Analysis

The SPSS22.0 and KANO model analysis methods were mainly adopted. Descriptive analysis in this research mainly selected frequency, percentage, mean and standard deviation for representation. Data statistical analysis methods in this research mainly included t-test, one-way analysis of variance, multiple linear regression analysis, etc.

The KANO model analysis method mainly analyzes the Kano attributes of the items of patients’ health education demand. According to the Kano quality attribute classification method, the characteristics of each item are classified as positive questions and reverse questions. The attribute with the highest proportion is the Kano attribute of this item. Among them, "A" represents attractive attributes, "M" represents must-be attributes, "O" represents one-dimensional attributes, "I" represents indifferent attributes, "R" represents reverse attributes, and "Q" represents questionable attributes. The attribution of the KANO model is shown in Table 1.

**Table 1.** Traditional Kano model categories.

| Dysfunctional<br>form | I like it<br>that way | It must<br>be that<br>way | I am<br>neutral | I can live<br>with it<br>that way | I dislike<br>it that<br>way |
| --- | --- | --- | --- | --- | --- |
| Functional form |  |  |  |  |  |
| I like it that way | Q | A | A | A | O |
| It must be that way | R | I | I | I | M |
| I am neutral | R | I | I | I | M |
| I can live with it that way | R | I | I | I | M |
| I dislike it that way | R | R | R | R | Q |

### Better-worse coefficient and draw the quadrant chart of importance - satisfaction

Since the traditional classification results are divided based on the maximum value, when the classification frequencies are the same or similar, the model cannot classify accurately. In this study, Better-worse coefficient analysis and four-quadrant graphs are used to describe satisfaction and importance, and the demand attributes are quantitatively analyzed. The better coefficient is the satisfaction coefficient. The calculation method is: (A+O)/(A+O+M+I). The closer it is to 1, the greater the impact of this item on patient satisfaction and the stronger the effect of improving satisfaction. The worse coefficient is the dissatisfaction coefficient, calculated as -1×(O+M)/(A+O+M+I). The closer it is to -1, the greater the impact of the item on the patient’s dissatisfaction and the stronger the effect of the reduction in satisfaction. The absolute value of the worse coefficient is the importance. Take the absolute value of the worse coefficient (importance) as the abscissa and the better coefficient (satisfaction) as the ordinate. With the mean values of importance and satisfaction as the dividing lines, the scatter plot is divided into four quadrants. The first, second, third and fourth quadrants respectively represent one-dimensional attribute (O), the attractive attribute (A), the indifferent attribute (I) and the must-be attribute (M).

### Ethical considerations

The study was carried out in accordance with the principles of the Declaration of Helsinki. This study was approved by the Medical Ethics Committee of West China Second Hospital of Sichuan University (No. 230 of the 2023 round of medical research approval).

### Participants’ characteristics and descriptive statistics

A total of 428 patients with HPV infection completed the questionnaire survey, with an average age of 40.49±10.56 years. Among them, the majority had a bachelor’s degree, accounting for 158 (36.9%), and 27.6% had a household monthly income of more than 10,000. More than 50% of the patients had cervical cancer screening once a year. Among the married patients, The majority (77.8%) have a cohabitation lifestyle with their spouses, and 83.9% of the patients have a sexual life frequency of 1 to 3 times a month. At present, more than 60% of unmarried patients have sexual partners. The lifestyle with their sexual partners is mostly cohabitation (49.3%), and the frequency of sexual life is mostly 1-3 times a month (59.7%). 86.7% of the patients had their first sexual life between the ages of 26 and 35. Among contraceptive measures, the usage rate of condoms reached 72.4%. 45.3% of the patients had been pregnant ≥3 times, and 58.9% of the patients had two or more sexual partners. The specific situation is shown in Table 2.

**Table 2.** Sample characteristics(N=428).

| Characteristics |  | n | % |
| --- | --- | --- | --- |
| Age (range: 19-75years) | Mean=40.49years<br>(SD=10.56) |  |  |
| Educational level | Below primary school | 5 | 1.2% |
|  | Primary school | 14 | 3.3% |
|  | Junior high school | 56 | 13.1% |
|  | High school or technical secondary school | 67 | 15.7% |
|  | Junior college | 94 | 22.0% |
|  | undergraduate | 158 | 36.9% |
|  | Master degree or above | 34 | 7.9% |
| Monthly household income (RMB) | $\leq 2000$ | 36 | 8.4% |
|  | 2001~4000 | 78 | 18.2% |
|  | 4001~6000 | 84 | 19.6% |
|  | 6001~8000 | 53 | 12.4% |
|  | 8001~10000 | 59 | 13.8% |
|  | >10000 | 118 | 27.6% |
| Cervical cancer screening frequency | Never | 70 | 16.4% |
|  | Once every 3 years or more | 44 | 10.3% |
|  | Every two years | 58 | 13.6% |
|  | Once a year | 220 | 51.4% |
|  | Multiple times a year | 36 | 8.4% |
| Marital status | Unmarried | 59 | 13.8% |
|  | Divorced | 47 | 11.0% |
|  | Widow | 6 | 1.4% |
|  | Married | 316 | 73.8% |
| Husband educational level | Below primary school | 3 | 0.9% |
|  | Primary school | 16 | 5.1% |
|  | Junior high school | 59 | 18.7% |
|  | High school or technical secondary school | 46 | 14.6% |
|  | Junior college | 67 | 21.2% |
|  | undergraduate | 107 | 33.9% |
|  | Master degree or above | 18 | 5.7% |
| lifestyle | cohabitation | 246 | 77.8% |
|  | Live apart | 56 | 17.7% |
|  | Other | 14 | 4.4% |
| Sexual frequency | 1-3 times a month | 234 | 83.9% |
|  | 1-3 times a week | 37 | 13.3% |
|  | 4-6 times a week | 5 | 1.8% |
|  | 7 or more times a week | 3 | 1.1% |
| Whether have a sexual partner | Yes | 69 | 61.6% |
|  | No | 43 | 38.4% |
| Sexual partner education level | Below primary school | 2 | 2.9% |
|  | Primary school | 1 | 1.4% |
|  | Junior high school | 6 | 8.7% |
|  | High school or technical secondary school | 11 | 15.9% |
|  | Junior college | 16 | 23.2% |
|  | undergraduate | 22 | 31.9% |
|  | Master degree or above | 11 | 15.9% |
| Lifestyle with sexual partners | cohabitation | 34 | 49.3% |
|  | Live apart | 31 | 44.9% |
|  | Other | 4 | 5.8% |
| Frequency of sexual intercourse with sexual partners | 1-3 times a month | 40 | 59.7% |
|  | 1-3 times a week | 16 | 23.9% |
|  | 4-6 times a week | 9 | 13.4% |
|  | 7 or more times a week | 2 | 3.0% |
| Age of first sexual intercourse | never | 0 | 0% |
|  | <16 | 0 | 0% |
|  | 16~25 | 11 | 2.6% |
|  | 26~35 | 371 | 86.7% |
|  | >35 | 46 | 10.7% |
| Contraceptive measure | None | 42 | 9.8% |
|  | condom | 310 | 72.4% |
|  | Oral contraceptive | 98 | 22.9% |
|  | Contraceptive ring | 104 | 24.3% |
|  | Other | 19 | 4.4% |
| Frequency of oral contraceptives | Less than once a month | 85 | 92.4% |
|  | 1-3 times a month | 5 | 5.4% |
|  | 3 or more times per month | 2 | 2.2% |
| Number of pregnancies | Never | 69 | 16.1% |
|  | 1 | 76 | 17.8% |
|  | 2 | 89 | 20.8% |
|  | ≥3 | 194 | 45.3% |
| Number of sexual partners | 1 | 176 | 41.1% |
|  | 2 | 106 | 24.8% |
|  | ≥3 | 146 | 34.1% |

### KANO model attribute classification of health education demand for patients with HPV infection

As shown in Table 3. The results of this study show that the health education demand of patients with HPV infection include 23 one-dimensional demand and 5 attractive demand, The Cronbach’s ɑ coefficient of the questionnaire was 0.952, the KMO value was 0.951, and the Bartlett sphericity test value was <0.001, which had good reliability and validity.

**Table 3.**
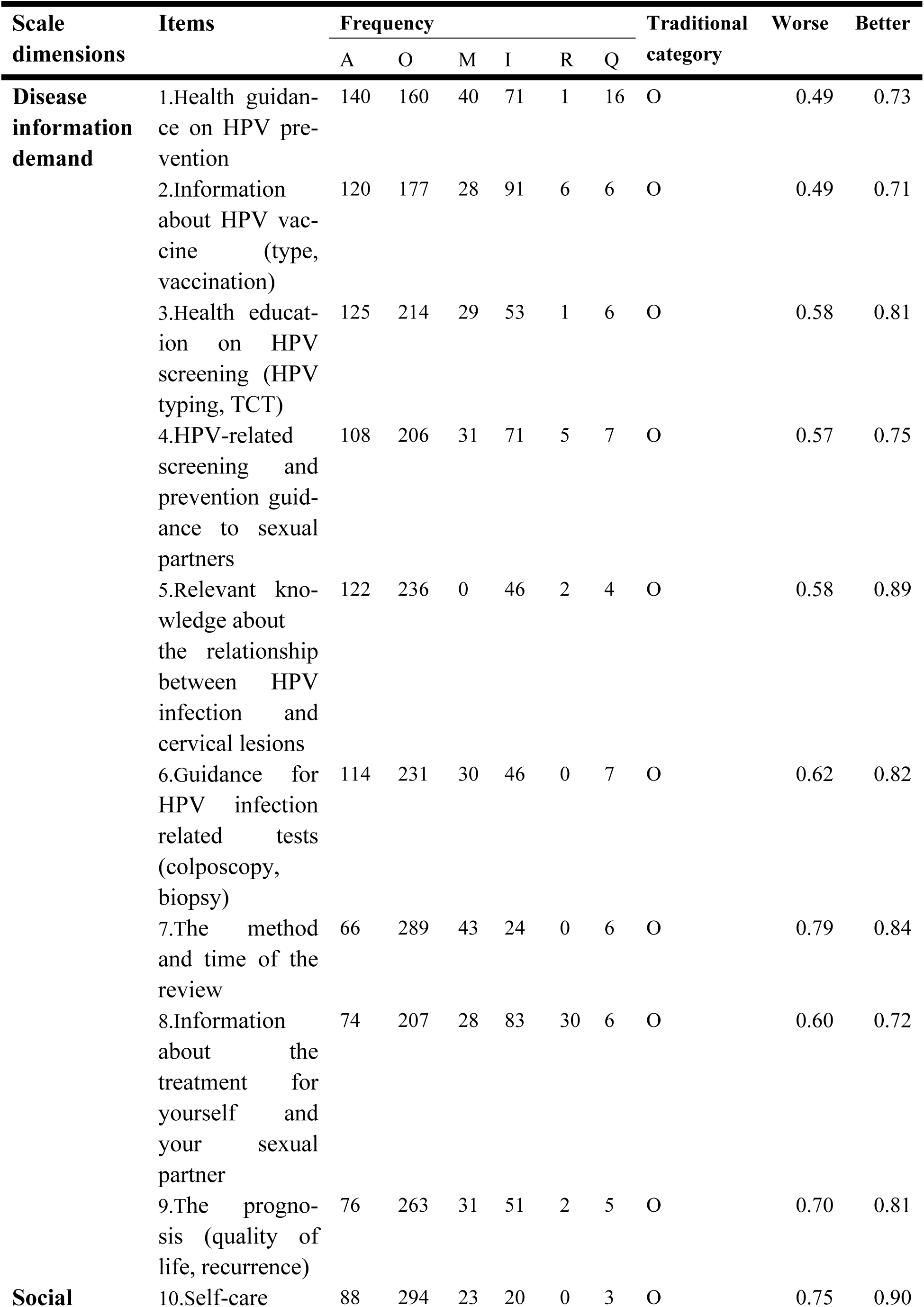
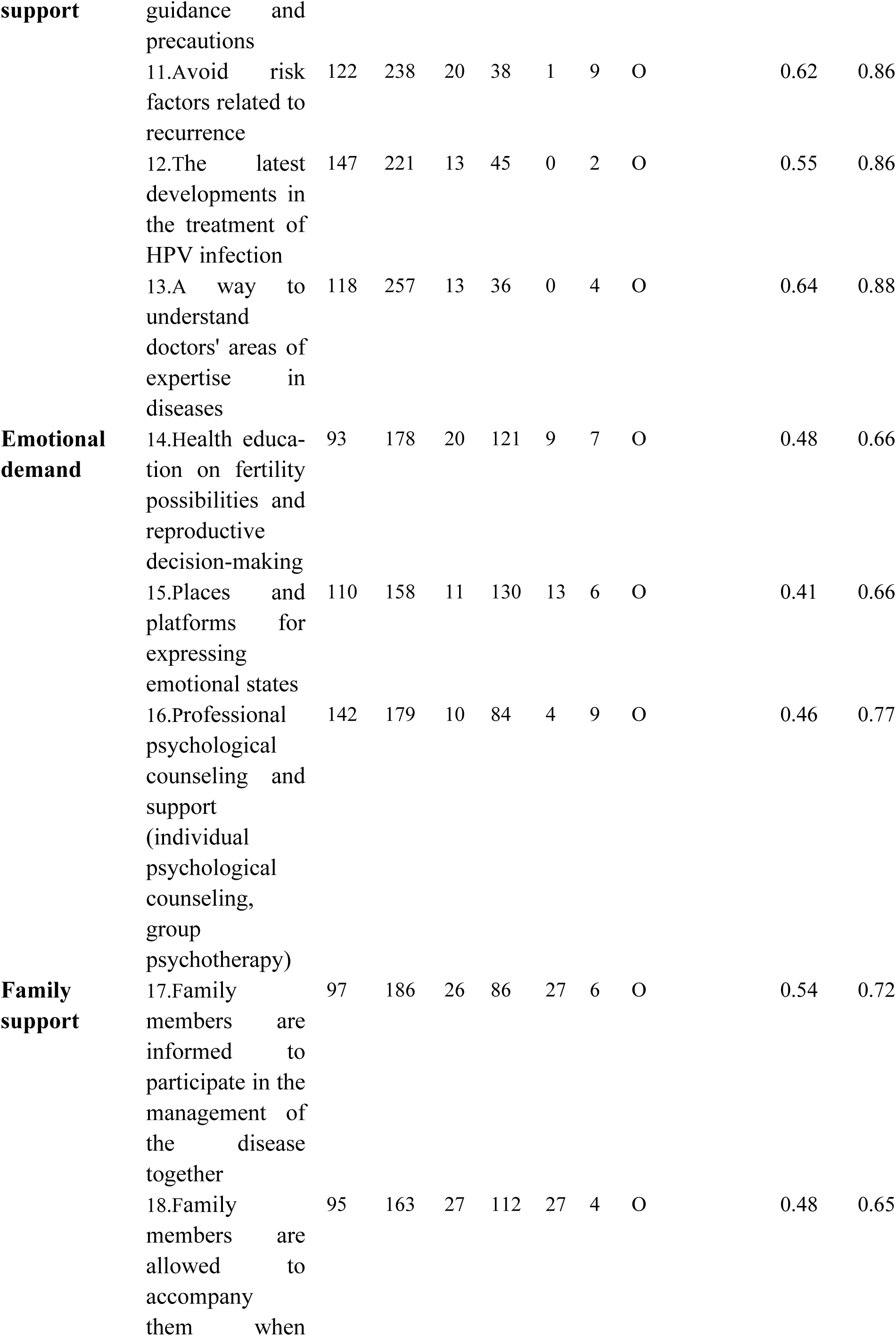
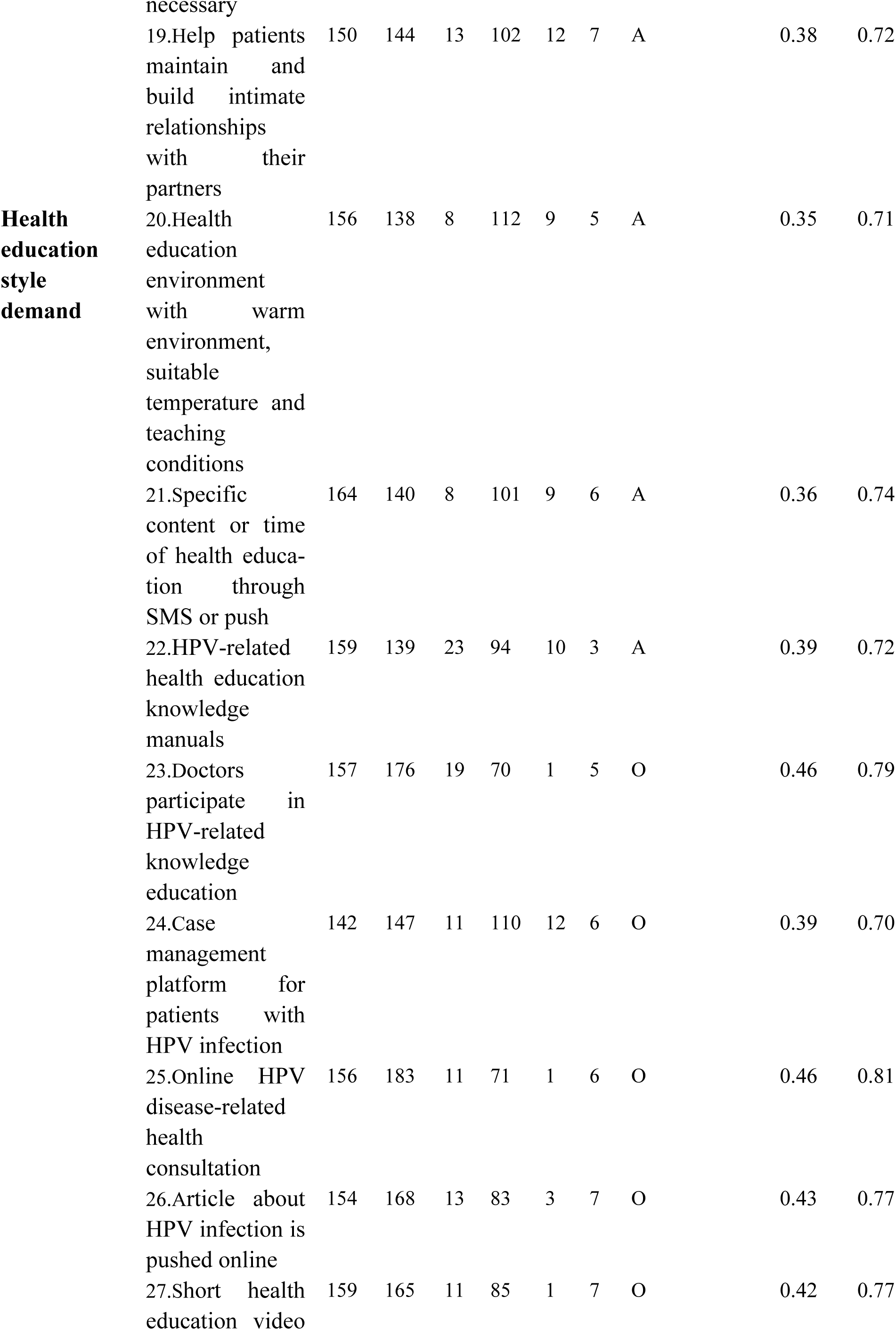
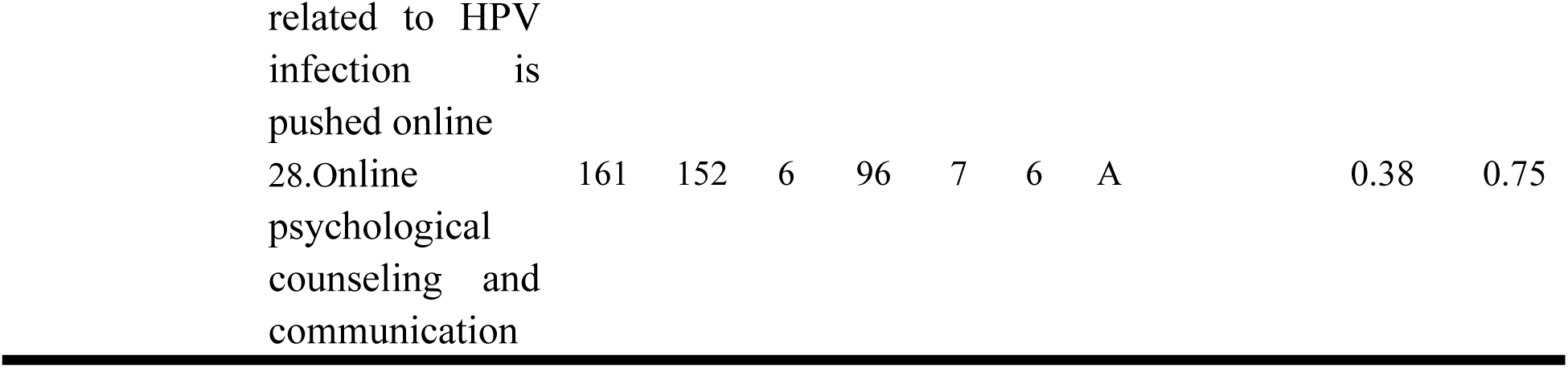
KANO Model Attributes of Health education demand for Patients with HPV infection.

### The importance of health education demand for patients with HPV infection - satisfaction

The importance and satisfaction of health education demand of patients with HPV infection were calculated according to the formula. A scatter plot was drawn with the importance as the horizontal axis and the satisfaction as the vertical axis. The scatter plot was divided into four quadrants respectively with the mean importance (0.513) and the mean satisfaction (0.768) of the 28 items as the dividing lines. Obtain the demand attributes of health education demand for patients with HPV infection: The attractive demand consists of 5 items, namely items 16, 23, 25, 26, and 27; The one-dimensional demand consist of 9 items, namely items 3, 5, 6, 7, 9, 10, 11, 12and 13. The must-be demand consist of three items, namely items 4, 8 and 17. The indifferent demand consist of 11 items, namely items 1, 2, 14, 15, 18, 19, 20, 21, 22, 24 and 28. (S1 Fig).

## Discussion

The analysis results of the demand attributes of this study show that the demands of patients with HPV infection include 23 one-dimensional demands and 5 attractive demands. Further through the better-worse coefficient analysis, it is found that the attractive demand contains 5 items, the one-dimensional demand contains 9 items, the must-be demand contains 3 items, and the indifferent demand contains 11 items. In contrast, It is more accurate to distinguish the demand of patients by using the better-worse coefficient. The traditional classification results are divided based on the maximum value, resulting in relatively absolute results. There may also be situations where there are two maximum values or frequencies close, making it difficult to determine the demand attributes. Classification is carried out through the Better-Worse coefficient, which to a certain extent compensates for the deficiencies of the traditional classification.

The Better-worse coefficient analysis shows that providing professional psychological counseling and support, doctors participating in the publicity and education of HPV-related knowledge, and conducting health education services online, including pushing articles and short videos, are attractive demands. Attractive demand is one of the demands that has the greatest impact on patient satisfaction. It is a demand attribute that exceeds patients’ expectations. The provision of services with such attributes can better improve patients’ satisfaction with continuous nursing services. At present, medical and health institutions seldom pay attention to the psychological changes of patients with HPV infection. However, a qualitative meta-integration result found that HPV infection can cause negative emotions in patients, affecting marital relationships and physical and mental health. HPV patients need disease knowledge, support from medical staff, and care from family and friends [15]. If medical staff can provide professional psychological counseling and support, it will greatly improve patients’ satisfaction. Meanwhile, patients have a strong demand for knowledge such as the early symptoms of cervical cancer (100.0%) and prevention (79.89%). They prefer to obtain health education through television/radio/the Internet (91.74%), health education prescriptions issued during diagnosis and treatment (83.47%), and face-to-face consultations (70.8%) [16]. If health education services for HPV can be provided online, the way for patients to obtain health education will be more convenient.

It is expected to provide disease information related to HPV, including how HPV is prevented, screened and prognostic, inform the latest developments in HPV infection treatment and offer ways to understand the doctor’s areas of expertise in the disease. One-dimensional demands are linear demands. Patient satisfaction is positively correlated with the degree to which their demand are met. When such demand are met, patient satisfaction often increases rapidly, and its importance is second only to must-be demand. Studies have found that the awareness rate of HPV-related knowledge in various regions of our country is 0%-45.81%, and the awareness rate is generally low. The awareness rate of HPV vaccines is only 8%-28.83%[17]. What women most desire to obtain is disease information related to HPV. This includes the transmission routes of HPV, prevention, detection, treatment, disease progression without treatment, and risk factors for cervical cancer. At the same time, women also want to know the duration of HPV infection, the regression situation, and the possibility of developing the disease [17]. In addition, there are many question records in online communities that also involve knowledge about HPV-related diseases, indicating that the public urgently demand the popularization of knowledge related to HPV infection. The content covers how to prevent HPV, the impact of HPV on pregnancy, etc. [18] Meanwhile, studies show that patients are eager to receive guidance and assistance from professional doctors and nurses to increase their understanding of disease-related knowledge [19]. The cognition of disease-related information is the basis of disease management, and such demands are also the ones that medical staff need to give priority to in the process of health education.

It is essential to provide guidance on HPV-related screening and prevention for sexual partners, offer relevant information on the treatment of both the individual and their sexual partners, and inform family members to jointly participate in the management of the disease. Such demands are the ones that need to be satisfied first and are of the highest importance. The main transmission route of HPV is sexual transmission. A meta-analysis including 65 global studies showed that the overall infection rate of any type of HPV in men worldwide was 31%, the infection rate of HR-HPV was 21%, and the most common HPV genotype for infection was HPV16(5%,95% CI:4% - 7%). The overall infection rate is relatively high [20]. On January 2, 2025, the National Medical Products Administration of China officially approved the quadrivalent HPV vaccine for use in men aged 9 to 26 and began vaccinations in multiple provinces and cities [21]. This indicates that HPV infection is no longer a disease that only targets women for intervention; intervention for men is equally important. Studies have found that spousal participatory health education can more effectively enhance women’s awareness of knowledge related to HPV infection and promote the joint formation of good preventive behaviors by both spouses [22]. In terms of social support, a qualitative study found that the respondents faced heavy psychological pressure, believing that HPV infection had a negative impact on their sexual relationships and family and social relationships [23]. They needed the support of their relatives and longed to be respected [24], and actively provided health education and guidance related to their sexual partners. Advocating the joint management of the disease by the family is the most important need for patients with HPV infection.

Regarding HPV prevention, vaccines, fertility decisions, as well as external conditions such as health education venues and environments, there were no differential demands in this study. Undifferentiated demand refer to demand without preferences, which are dispensable for patients. Since 2018, China has started HPV vaccination for women. The vaccination rate of HPV among women aged 9 to 45 has been increasing year by year. The public’s understanding of cervical cancer vaccines has gradually deepened. Compared with other demands, the importance of this demand is relatively low, which also proves that health education on HPV prevention and vaccines has been carried out well in recent years. In terms of fertility decision-making, since the average age of the subjects investigated in this study was 40.49 years old and the possibility of having another child was relatively low, such demands were not necessary. However, some researchers analyzed the public’s HPV-related question records through online platforms, and there were still many question records involving the impact of HPV infection on pregnancy [18]. Therefore, such demands should also take into account the patient’s age and actual reproductive demand. Regarding the environment for health education, as well as the provision of external conditions such as places for expressing emotions, online psychological counseling, and the establishment of case management platforms, patients may pay more attention to the health education provided by medical staff who are professional enough and have good communication skills [25], rather than external conditions. As for allowing family members to accompany patients during medical consultations when necessary to help them maintain and establish an intimate relationship with their partners, what patients need more is the joint participation of their spouses in disease management. Spousal participatory health education can better promote the formation of good preventive behaviors between the couple [22].

## Limitation

The population in this study is only focused on one region, which is limited and the representative sample is insufficient.

## Conclusion

This study analyzed the health education demand of patients with HPV infection based on the HANO model and found that the demand related to sexual partners were of the highest importance. The psychological demand of patients with HPV infection should be taken seriously. The cognition of disease-related information is the basis of disease management, while the demand for related health education methods belong to undifferentiated demand.

## Data Availability

All relevant data are within the manuscript and its Supporting Information files.

## Acknowledgments

We are very grateful to the Patient Follow-up Center of West China Second Hospital of Sichuan University for its contribution to the collection of questionnaires. We would like to thank Mrs.Yang for valuable contributions to this research, including collaboration in experiments and data collection.

## Supporting information

**S1 Fig.**
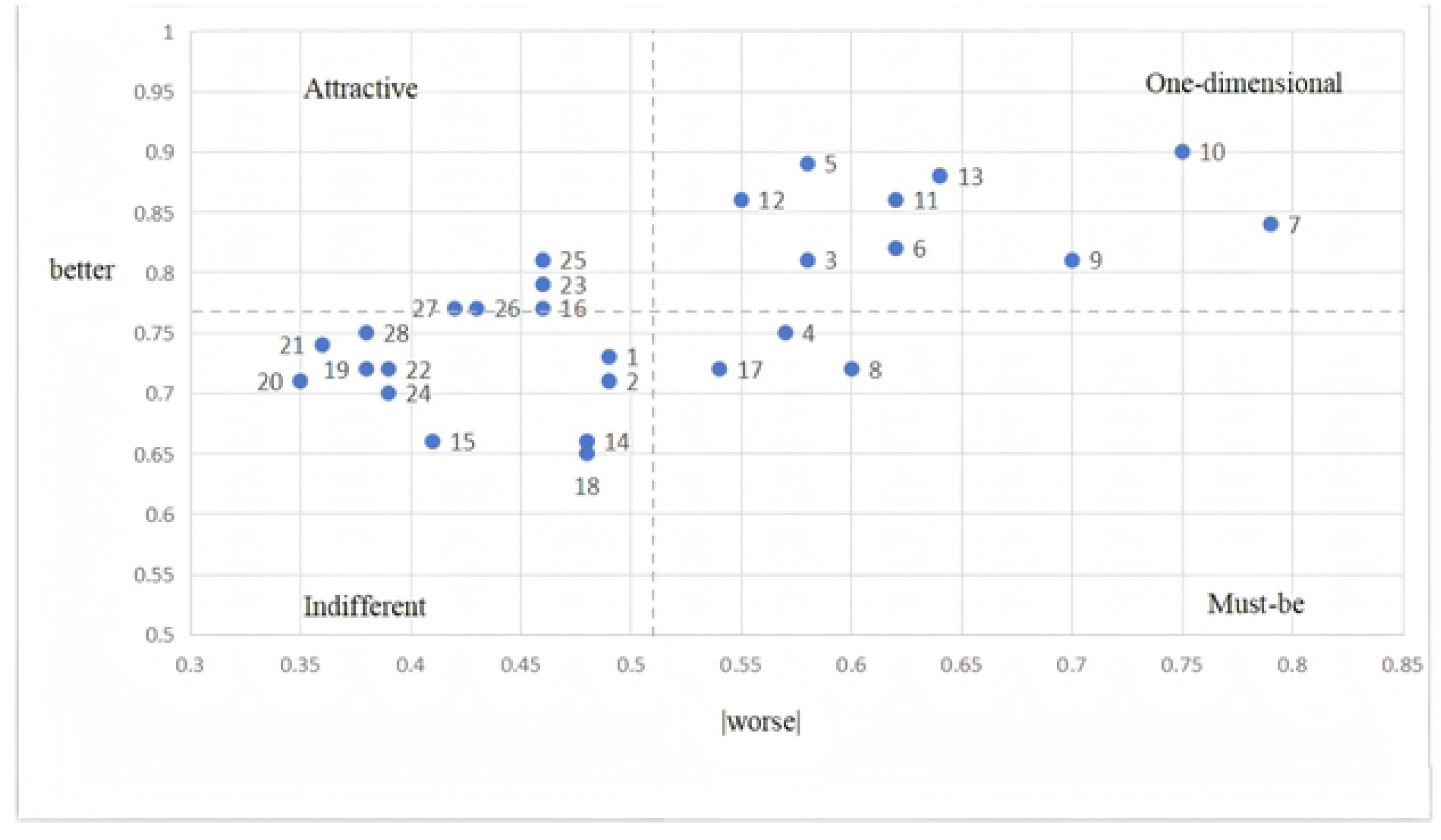
Attributes on the better-worse plot.

**The data**

**The health education demand scale for HPV infected.**

